# Discordant Associations of Food- and Nutrient-Based Dietary Inflammatory Scores With Cognitive Performance in Older Adults: A Cross-Sectional Study

**DOI:** 10.64898/2026.08.02.26359519

**Authors:** Jian Zheng, Jiayu Du, Zijie Liu, Shuyue Jin, Kaiguo Tian, Jingxuan Wang, Qi Zhang

**Affiliations:** College of Pharmacy, Shandong Provincial Engineering Laboratory of Novel Pharmaceutical Excipients Sustained and Controlled Release Preparations, Dezhou University, Dezhou 253023, China; Dezhou Qihe County Cultural Tourism Bureau, Dezhou 251100, China

**Keywords:** Food Inflammation Score for Individuals, Dietary Inflammatory Index, cognitive performance, older adults, NHANES, complex survey

## Abstract

**Background & aims:** Food- and nutrient-based inflammatory scores may capture different dietary features. We compared the Food Inflammation Score for Individuals (FISI) and Dietary Inflammatory Index (DII) in relation to cognitive performance and incremental discrimination.

**Methods:** This cross-sectional analysis included 2,455 adults aged ≥60 years in NHANES 2011-2014 with four cognitive tests and two reliable 24-h recalls. Daily FISI summed [(food FII per 100 g) × grams/100] across foods; two days were averaged. DII used 33 parameters averaged across the same days. Survey-weighted linear and logistic models incorporated strata, primary sampling units, and WTDR2D/2. Discrimination used survey-weighted AUROCs and 500 stratified primary-sampling-unit bootstrap replicates.

**Results:** After full adjustment, a 1-SD higher DII was associated with a 0.091 lower global cognitive Z-score (β = −0.091; 95% CI, −0.143 to −0.040; P = 0.005), whereas FISI was not (β = −0.019; 95% CI, −0.064 to 0.026; P = 0.341). The DII quartile trend was significant (P = 0.007), but nominal domain associations did not survive false-discovery-rate correction. Neither score was associated with low cognitive performance. The base AUROC was 0.8242; adding FISI changed it by <0.0001 (95% bootstrap CI, −0.0002 to 0.0007), and adding DII changed it by 0.0005 (−0.0003 to 0.0021).

**Conclusions:** Higher DII, but not FISI, was associated with lower global cognitive performance. Neither score materially improved discrimination for low cognitive performance. The two scores should not be assumed equivalent or interchangeable.

## 1. Introduction

Cognitive decline and dementia are major causes of disability and dependency in later life. Diet is a potentially modifiable factor, but its role is difficult to isolate because foods, nutrients, health behaviors, and cardiometabolic conditions are interrelated [1]. Chronic low-grade inflammation may link dietary exposures to cognitive aging. A systematic review in older adults found generally adverse associations between dietary inflammatory potential and cognition but substantial variation in dietary assessment, index construction, eligibility criteria, covariate control, and study design [2]. A recent prospective-cohort meta-analysis likewise reported greater cognitive-impairment risk with more pro-inflammatory diets and emphasized standardized exposure measurement [3].

The Dietary Inflammatory Index (DII) is a literature-derived, population-referenced score that integrates the inflammatory weights of multiple dietary parameters [4]. It has been validated against circulating C-reactive protein [5] and is widely used in nutritional epidemiology. However, the DII is calculated at the individual nutrient or food-parameter level and its implementation depends on the parameters available in a given dietary instrument. A newer Food Inflammation Index (FII) assigns an inflammatory value to each food per 100 g from its composition, and the corresponding Food Inflammation Score for Individuals (FISI) aggregates the FII values of foods actually consumed [6]. This food-level framework may preserve within-food information that is not represented by a nutrient-sum score, but FISI remains comparatively new and should not be presumed to reproduce the DII construct.

Several cross-sectional analyses of the 2011-2014 National Health and Nutrition Examination Survey (NHANES) have linked higher DII values to poorer performance on one or more cognitive tests [7–9]. Prospective studies have also associated pro-inflammatory diets with incident dementia or later cognitive performance [10,11], although other work has reported null or mixed associations with global cognition and brain imaging measures [12]. Long-term and community-based studies in other populations have likewise produced inverse associations of varying magnitude [13,14], and a broader meta-analysis has documented heterogeneity across cognitive outcomes [15]. Differences across studies may reflect population, dietary instrument, exposure window, score implementation, outcome definition, and adjustment strategy. Direct comparison of food- and nutrient-based inflammatory scores within the same survey design may therefore clarify whether they yield similar epidemiological signals.

We compared two-day FISI and a 33-parameter two-day DII in adults aged ≥60 years from NHANES 2011-2014. The primary objective was to estimate their associations with a standardized global cognitive score under the same complex-survey framework. Secondary objectives were to examine cognitive domains, low cognitive performance, dose-response patterns, robustness across sensitivity analyses, and incremental discrimination beyond demographic, socioeconomic, behavioral, dietary, and cardiometabolic covariates. We did not prespecify or test an equivalence margin; consequently, similarity or non-significance was not interpreted as evidence that the scores were equivalent.

## 2. Materials and methods

### 2.1 Study design and population

NHANES is a continuous, nationally representative survey of the civilian, non-institutionalized population of the United States. It uses a complex, multistage probability design that requires incorporation of sample weights, strata, and primary sampling units in analysis [16]. We combined the 2011-2012 and 2013-2014 cycles because the same cognitive battery was administered to participants aged ≥60 years.

The National Center for Health Statistics Ethics Review Board approved NHANES 2011-2012 under Protocol #2011-17 and NHANES 2013-2014 as a continuation of that protocol; participants provided written informed consent [17]. The present study used deidentified public-use data and involved no additional participant contact.

Starting with 19,931 participants, we sequentially retained those aged ≥60 years (n = 3,632), with complete results for all four cognitive tests (n = 2,934), with two reliable dietary recalls (n = 2,524), with a positive two-day dietary weight and complete survey-design variables (n = 2,524), and with calculable two-day FISI and 33-parameter DII values (n = 2,524). We then excluded 16 participants with implausible sex-specific two-day mean energy intake (<500 or >5,000 kcal/day for men; <500 or >3,500 kcal/day for women) and 53 with body mass index (BMI) outside 15-50 kg/m² or missing core demographic data. The final analytic sample comprised 2,455 participants (Figure 1; Supplementary Table S1).

**Figure 1.**
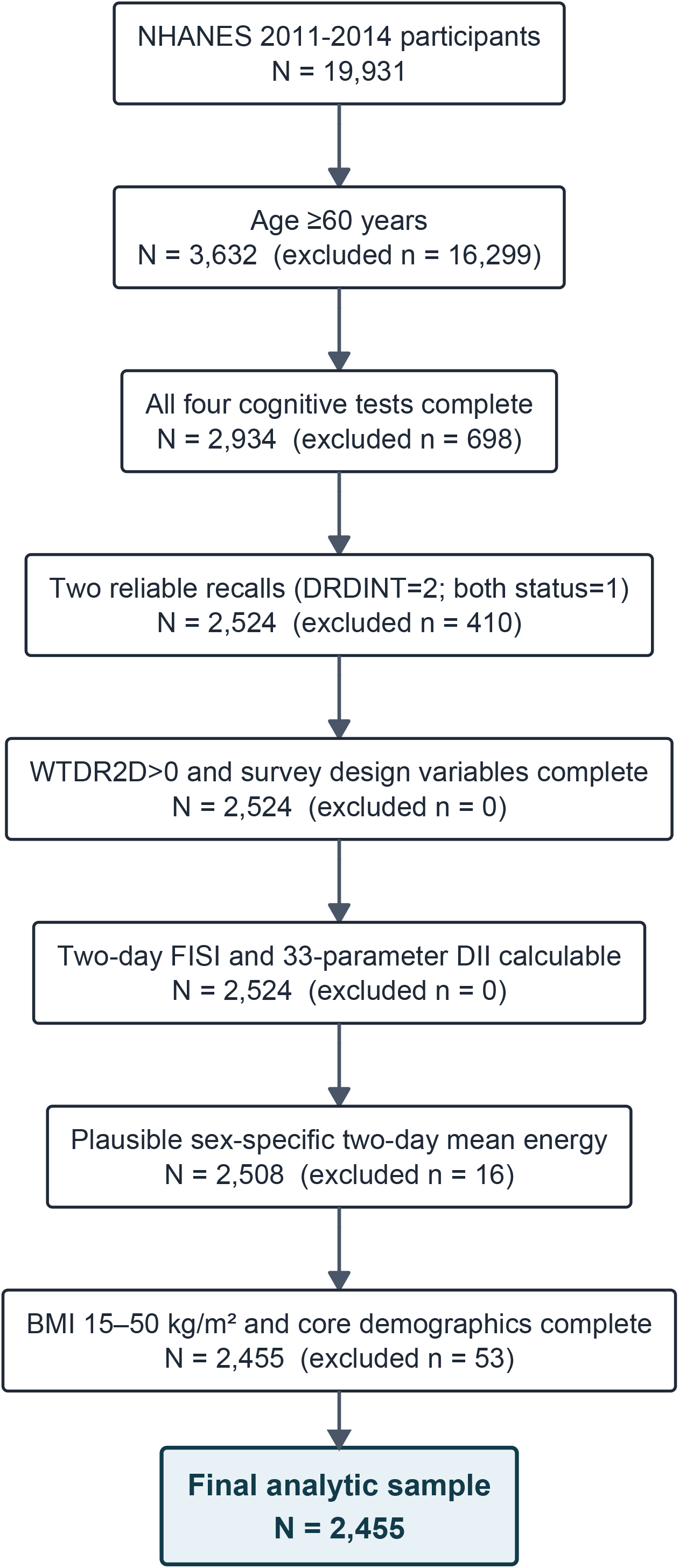
Participant selection. Sequential selection of the frozen v3 analytic sample from NHANES 2011–2014. BMI, body mass index; DII, Dietary Inflammatory Index; FISI, Food Inflammation Score for Individuals; NHANES, National Health and Nutrition Examination Survey. Preferred final width: two columns.

### 2.2 Cognitive outcomes

NHANES administered the Consortium to Establish a Registry for Alzheimer’s Disease (CERAD) Word Learning test, the Animal Fluency Test, and the Digit Symbol Substitution Test (DSST) [18]. The CERAD immediate-recall score was the sum of three learning trials; the delayed-recall score was analyzed separately. Animal Fluency assessed semantic verbal fluency, and DSST assessed processing speed, sustained attention, and working memory. These tests characterize cognitive performance but do not establish a clinical diagnosis [18–20].

Within the survey-weighted analytic sample, each of the four raw scores was standardized to a Z-score. Their mean was then re-standardized to form the global cognitive Z-score, with higher values indicating better performance. The primary outcome was continuous global cognitive performance. Secondary continuous outcomes were the four domain-specific Z-scores. Low cognitive performance was defined as a global Z-score at or below its survey-weighted 25th percentile (−0.697), and was explicitly treated as a performance threshold rather than a diagnosis.

### 2.3 Dietary recalls and two-day exposure window

Dietary intake was measured with two 24-h recalls. The first was collected in person in the mobile examination center and the second by telephone on a later non-consecutive day. We included only participants for whom both recall statuses were reliable, the number-of-days indicator was DRDINT = 2, and the two-day dietary weight WTDR2D was positive [21]. Energy and all dietary parameters were averaged across Day 1 and Day 2 so that the covariate and exposure windows were aligned.

### 2.4 Food Inflammation Index and Food Inflammation Score for Individuals

FII was used only for the food-level index expressed per 100 g. The individual exposure was named FISI. Following the original FII framework [6], each unique food code within each NHANES cycle was assigned a raw food-level value:

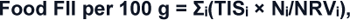

where TISᵢ is the total inflammatory score for component *i*, Nᵢ is the amount of that component per 100 g of food, and NRVᵢ is its reference value. Thirty-four components came directly from the NHANES individual-food nutrient files. Five flavonoid subclasses— flavan-3-ols, flavanones, flavones, flavonols, and isoflavones—were linked by USDA food code from the Database of Flavonoid Values for USDA Food Codes 2007-2010 [22] (Supplementary Table S11). Unmatched flavonoid values were set to zero. Across repeated occurrences of a food code within a cycle, component values per 100 g were summarized by the median. Raw food-level FII values were winsorized at the cycle-specific 1st and 99th percentiles to reduce the influence of extreme food-composition values.

For each participant and recall day:

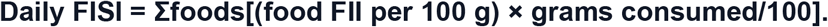

The primary FISI was the mean of Day 1 and Day 2. It was not divided by the total grams of food consumed and was not normalized to a 1-100 scale. As a separate aggregation check, daily and two-day FISI were re-aggregated food by food from the stored food-level FII values for ten participants spanning the FISI distribution; the maximum absolute difference between the production and re-aggregated two-day values was 1.78 × 10⁻¹⁵ (Supplementary Table S10).

### 2.5 Dietary Inflammatory Index

DII was calculated according to the published population-referenced algorithm [4]. Of the 45 candidate parameters, 33 were available in both cycles and on both recall days (Supplementary Table S9). For each participant, parameters were averaged across the two days. Each two-day mean was standardized using the published global mean and standard deviation, transformed to a centerd percentile score [2Φ(z) - 1], multiplied by its literature-derived inflammatory effect score, and summed. Higher DII values indicate greater pro-inflammatory potential. All included participants had values for all 33 implemented parameters.

### 2.6 Covariates

Covariates were selected before the v3 rerun based on prior literature and availability in both cycles. They comprised age; sex; race/ethnicity; education (<high school, high school, >high school, or unknown); marital status (partnered, not partnered, or unknown); poverty-income ratio (PIR); smoking (never, former, current, or unknown); alcohol intake (g/day); recreational physical activity (active, inactive, or unknown); BMI; two-day mean energy intake; hypertension; diabetes; and hyperlipidemia.

Physical activity was coded as active when a participant reported vigorous or moderate recreational activity and inactive when both were denied; no metabolic-equivalent minutes were derived. Diabetes was defined by self-report or glycated hemoglobin ≥6.5%. Hypertension was based on self-report. Hyperlipidemia was defined by self-reported high cholesterol, total cholesterol ≥240 mg/dL, or lipid-lowering medication use.

PIR was missing for 186 participants and was imputed with the survey-weighted median; a missingness indicator was included. Alcohol intake was unavailable for 1,063 participants, was set to zero for model stability, and was accompanied by a missingness indicator. Unknown categories retained participants with missing categorical covariates. The complete-case sensitivity analysis excluded all such imputed or unknown values (Supplementary Table S12).

### 2.7 Statistical analysis

Analyses followed STROBE reporting recommendations [23]. All estimates incorporated SDMVSTRA, SDMVPSU, and the four-year dietary weight WTDR2D/2. Continuous variables are reported as survey-weighted means and standard deviations or weighted quantiles; categorical variables are reported as weighted percentages. FISI and DII were standardized using their survey-weighted means and standard deviations. Survey-weighted quartiles were also constructed for each score. The weighted Spearman correlation was reported descriptively.

Survey-weighted linear regression estimated differences in global or domain-specific cognitive Z-scores per 1-SD higher exposure. Model 1 adjusted for age and sex. Model 2 additionally adjusted for race/ethnicity, education, marital status, PIR, and PIR missingness. Model 3 additionally adjusted for smoking, alcohol intake and missingness, physical activity, BMI, two-day energy intake, hypertension, diabetes, and hyperlipidemia. Quartile models used Q1 as the reference; trends were tested by entering the ordinal quartile variable continuously. Survey-weighted quasibinomial regression estimated odds ratios (ORs) for low cognitive performance. The domain-specific P values were corrected with the Benjamini-Hochberg false discovery rate (FDR).

Restricted cubic splines with knots at the survey-weighted 5th, 35th, 65th, and 95th percentiles evaluated non-linearity [24]. A mutually adjusted Model 3 included FISI and DII simultaneously. Prespecified interaction models assessed diabetes, obesity, and NHANES cycle; their six P values were FDR-corrected. Sensitivity analyses used complete covariate cases, FISI per 1,000 kcal, exclusion of self-reported stroke or coronary heart disease, flavonoid gram coverage ≥95%, un-winsorized food-level FII, separate survey cycles, and alternative 20th- and 30th-percentile thresholds for low cognitive performance.

For exploratory discrimination, a base quasibinomial model included all Model 3 covariates. We compared its survey-weighted AUROC with models adding FISI or DII. Confidence intervals and two-sided P values were obtained from 500 bootstrap replicates that resampled primary sampling units with replacement within strata and refitted each model. AUROC differences were interpreted as discrimination changes, not as tests of score equivalence or clinical utility [25]. Historical unweighted DeLong, net reclassification improvement, and integrated discrimination improvement analyses were not used.

All tests were two-sided. P < 0.05 was considered statistically significant for the primary analysis; multiplicity-adjusted results are reported for domain and interaction families. Analyses were performed in R version 4.6.1 using the survey package version 4.5. The frozen v3 scripts parsed without error, all five core error logs were empty, and no frozen primary model changed the 2,455-participant analytic sample.

## 3. Results

### 3.1 Participant and exposure characteristics

The 2,455 participants represented a four-year weighted population of approximately 53.6 million older adults. The survey-weighted mean age was 69.1 years (SD 6.6), 53.5% were women, and 78.7% were non-Hispanic White (Table 1). Participants in higher FISI quartiles were older and more often women; several socioeconomic and behavioral characteristics also differed across quartiles.

**Table 1.**
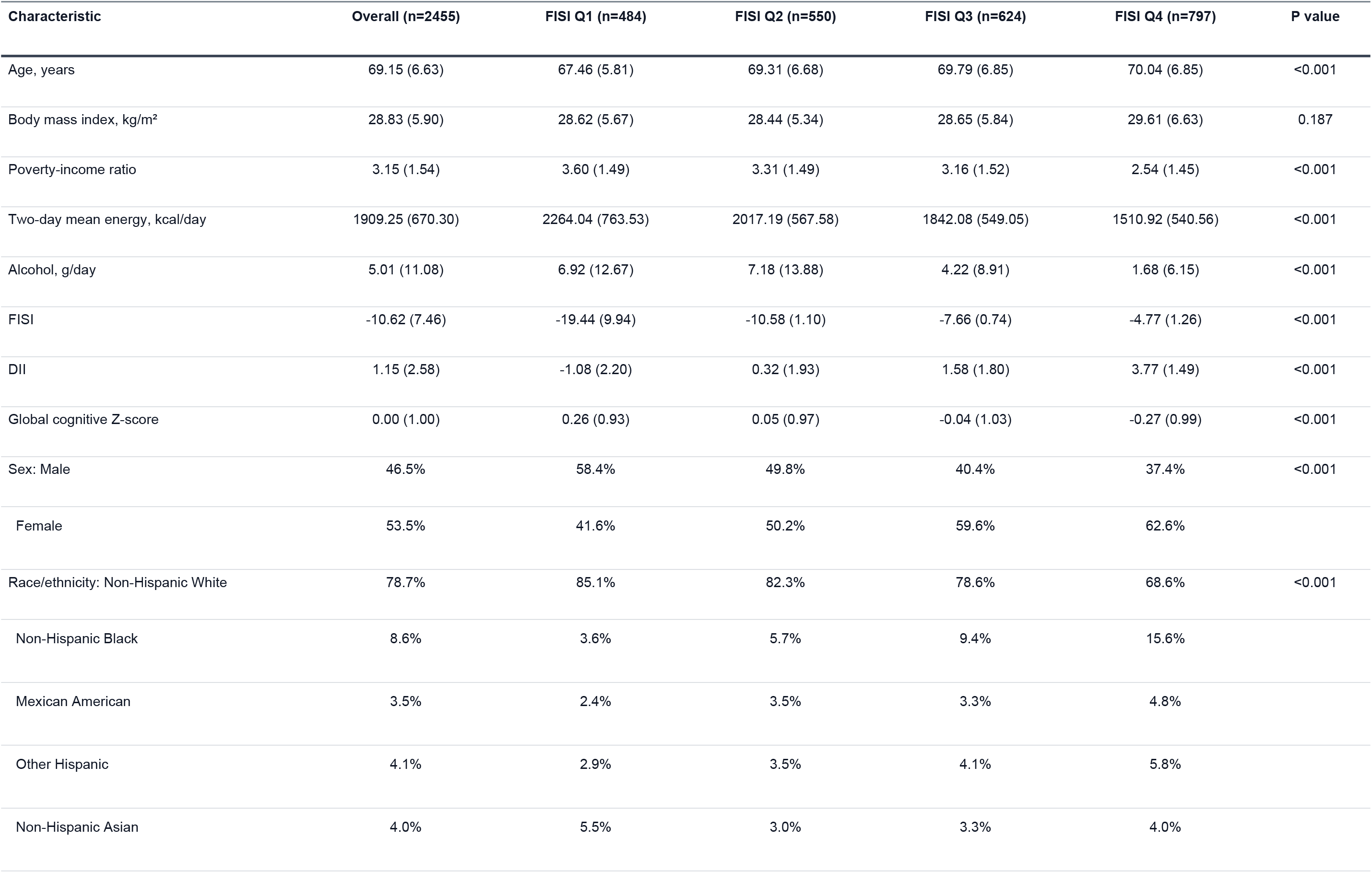
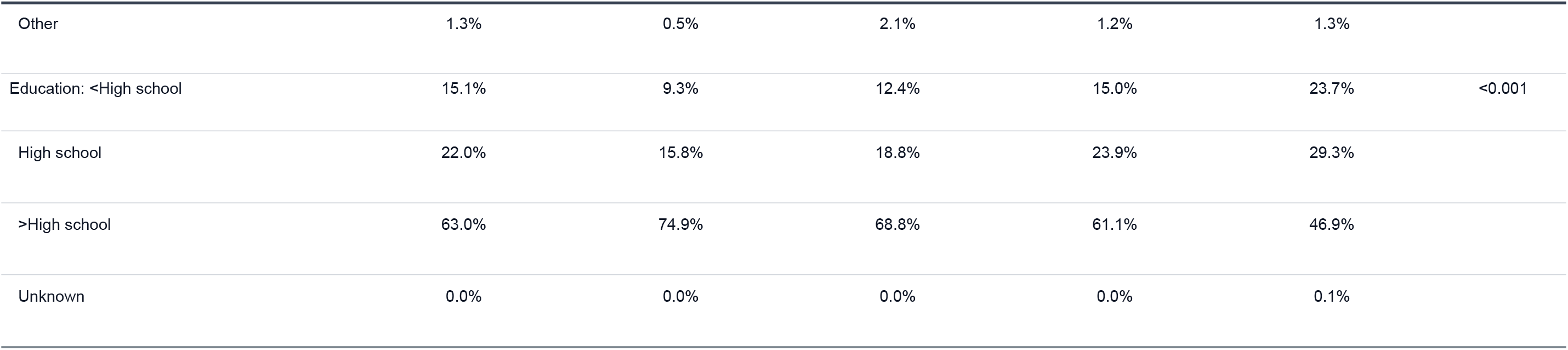
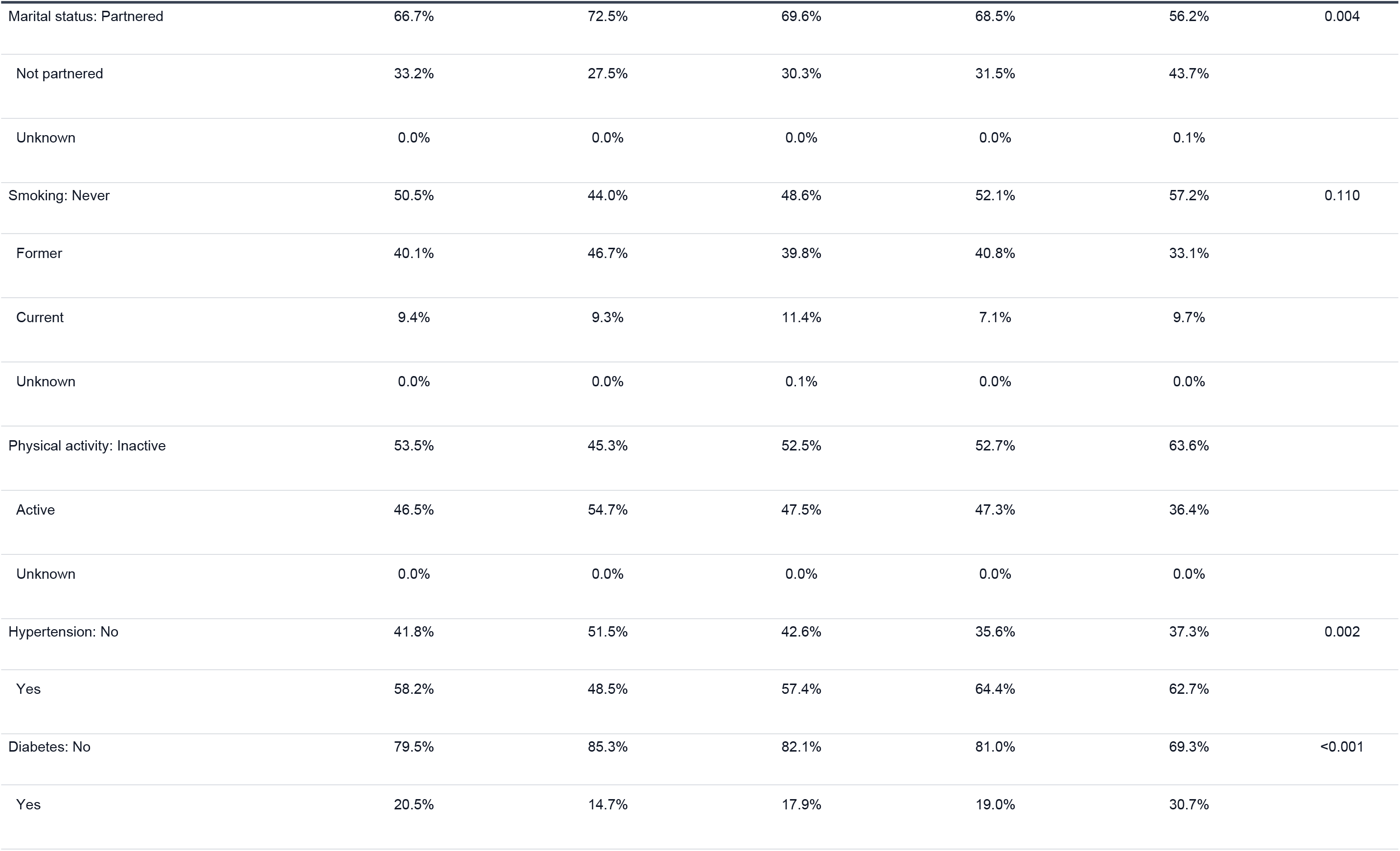
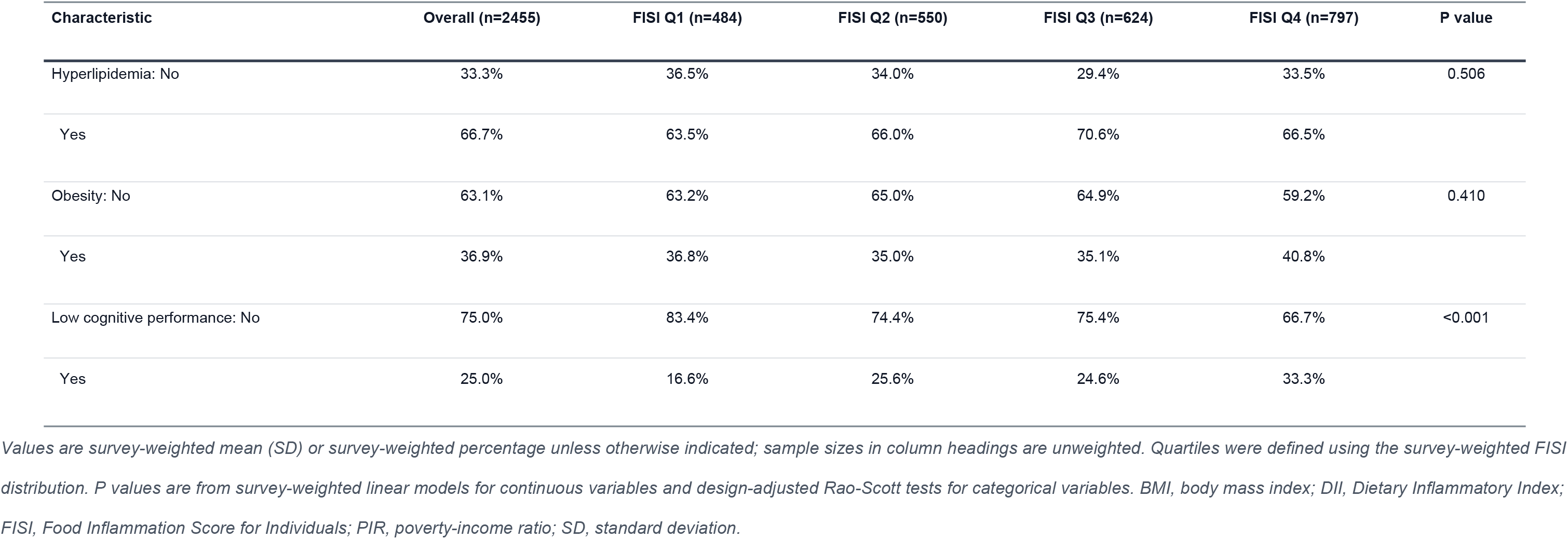
Survey-weighted participant characteristics overall and by FISI quartile.

| Characteristic | Overall (n=2455) | FISI Q1 (n=484) | FISI Q2 (n=550) | FISI Q3 (n=624) | FISI Q4 (n=797) | P value |
| --- | --- | --- | --- | --- | --- | --- |
| Age, years | 69.15 (6.63) | 67.46 (5.81) | 69.31 (6.68) | 69.79 (6.85) | 70.04 (6.85) | <0.001 |
| Body mass index, kg/m<sup>2</sup> | 28.83 (5.90) | 28.62 (5.67) | 28.44 (5.34) | 28.65 (5.84) | 29.61 (6.63) | 0.187 |
| Poverty-income ratio | 3.15 (1.54) | 3.60 (1.49) | 3.31 (1.49) | 3.16 (1.52) | 2.54 (1.45) | <0.001 |
| Two-day mean energy, kcal/day | 1909.25 (670.30) | 2264.04 (763.53) | 2017.19 (567.58) | 1842.08 (549.05) | 1510.92 (540.56) | <0.001 |
| Alcohol, g/day | 5.01 (11.08) | 6.92 (12.67) | 7.18 (13.88) | 4.22 (8.91) | 1.68 (6.15) | <0.001 |
| FISI | -10.62 (7.46) | -19.44 (9.94) | -10.58 (1.10) | -7.66 (0.74) | -4.77 (1.26) | <0.001 |
| DII | 1.15 (2.58) | -1.08 (2.20) | 0.32 (1.93) | 1.58 (1.80) | 3.77 (1.49) | <0.001 |
| Global cognitive Z-score | 0.00 (1.00) | 0.26 (0.93) | 0.05 (0.97) | -0.04 (1.03) | -0.27 (0.99) | <0.001 |
| Sex: Male | 46.5% | 58.4% | 49.8% | 40.4% | 37.4% | <0.001 |
| Female | 53.5% | 41.6% | 50.2% | 59.6% | 62.6% |  |
| Race/ethnicity: Non-Hispanic White | 78.7% | 85.1% | 82.3% | 78.6% | 68.6% | <0.001 |
| Non-Hispanic Black | 8.6% | 3.6% | 5.7% | 9.4% | 15.6% |  |
| Mexican American | 3.5% | 2.4% | 3.5% | 3.3% | 4.8% |  |
| Other Hispanic | 4.1% | 2.9% | 3.5% | 4.1% | 5.8% |  |
| Non-Hispanic Asian | 4.0% | 5.5% | 3.0% | 3.3% | 4.0% |  |
| Other | 1.3% | 0.5% | 2.1% | 1.2% | 1.3% |  |
| Education: <High school | 15.1% | 9.3% | 12.4% | 15.0% | 23.7% | <0.001 |
| High school | 22.0% | 15.8% | 18.8% | 23.9% | 29.3% |  |
| >High school | 63.0% | 74.9% | 68.8% | 61.1% | 46.9% |  |
| Unknown | 0.0% | 0.0% | 0.0% | 0.0% | 0.1% |  |

Table 1 (continued). Survey-weighted participant characteristics overall and by FISI quartile
| Characteristic | Overall (n=2455) | FISI Q1 (n=484) | FISI Q2 (n=550) | FISI Q3 (n=624) | FISI Q4 (n=797) | P value |
| --- | --- | --- | --- | --- | --- | --- |
| Marital status: Partnered | 66.7% | 72.5% | 69.6% | 68.5% | 56.2% | 0.004 |
| Not partnered | 33.2% | 27.5% | 30.3% | 31.5% | 43.7% |  |
| Unknown | 0.0% | 0.0% | 0.0% | 0.0% | 0.1% |  |
| Smoking: Never | 50.5% | 44.0% | 48.6% | 52.1% | 57.2% | 0.110 |
| Former | 40.1% | 46.7% | 39.8% | 40.8% | 33.1% |  |
| Current | 9.4% | 9.3% | 11.4% | 7.1% | 9.7% |  |
| Unknown | 0.0% | 0.0% | 0.1% | 0.0% | 0.0% |  |
| Physical activity: Inactive | 53.5% | 45.3% | 52.5% | 52.7% | 63.6% |  |
| Active | 46.5% | 54.7% | 47.5% | 47.3% | 36.4% |  |
| Unknown | 0.0% | 0.0% | 0.0% | 0.0% | 0.0% |  |
| Hypertension: No | 41.8% | 51.5% | 42.6% | 35.6% | 37.3% | 0.002 |
| Yes | 58.2% | 48.5% | 57.4% | 64.4% | 62.7% |  |
| Diabetes: No | 79.5% | 85.3% | 82.1% | 81.0% | 69.3% | <0.001 |
| Yes | 20.5% | 14.7% | 17.9% | 19.0% | 30.7% |  |
| Hyperlipidemia: No | 33.3% | 36.5% | 34.0% | 29.4% | 33.5% | 0.506 |
| Yes | 66.7% | 63.5% | 66.0% | 70.6% | 66.5% |  |
| Obesity: No | 63.1% | 63.2% | 65.0% | 64.9% | 59.2% | 0.410 |
| Yes | 36.9% | 36.8% | 35.0% | 35.1% | 40.8% |  |
| Low cognitive performance: No | 75.0% | 83.4% | 74.4% | 75.4% | 66.7% | <0.001 |
| Yes | 25.0% | 16.6% | 25.6% | 24.6% | 33.3% |  |

Values are survey-weighted mean (SD) or survey-weighted percentage unless otherwise indicated; sample sizes in column headings are unweighted. Quartiles were defined using the survey-weighted FISI distribution. P values are from survey-weighted linear models for continuous variables and design-adjusted Rao-Scott tests for categorical variables. BMI, body mass index; DII, Dietary Inflammatory Index; FISI, Food Inflammation Score for Individuals; PIR, poverty-income ratio; SD, standard deviation.

The weighted mean FISI was −10.62 (SD 7.46), with a median of −8.95 (interquartile range, −12.89 to −6.34). The weighted mean DII was 1.15 (SD 2.58), with a median of 1.25 (interquartile range, −0.65 to 3.07). FISI and DII had a weighted Spearman correlation of 0.726. The USDA flavonoid linkage covered 88.3% of food records and 89.8% of consumed grams overall; gram-weighted coverage was 93.3% in 2011-2012 and 86.3% in 2013-2014 (Supplementary Table S2).

### 3.2 Primary continuous-outcome models

In Model 1, each 1-SD higher FISI was associated with a 0.123 lower global cognitive Z-score (95% CI, −0.199 to −0.048; P = 0.002). The estimate attenuated after socioeconomic adjustment (Model 2: β = −0.052; 95% CI, −0.107 to 0.003; P = 0.062) and was not significant in Model 3 (β = −0.019; 95% CI, −0.064 to 0.026; P = 0.341) (Table 2).

**Table 2.** Survey-weighted associations of FISI and DII with cognitive performance.

| Panel | Exposure | Contrast | $\beta$ or OR (95% CI) | P value | n |
| --- | --- | --- | --- | --- | --- |
| A. Continuous global cognitive performance | FISI | Model 1: per 1-SD higher score | -0.123 (-0.199 to -0.048) | 0.002 | 2455 |
|  | FISI | Model 2: per 1-SD higher score | -0.052 (-0.107 to 0.003) | 0.062 | 2455 |
|  | FISI | Model 3: per 1-SD higher score | -0.019 (-0.064 to 0.026) | 0.341 | 2455 |
|  | DII | Model 1: per 1-SD higher score | -0.230 (-0.275 to -0.185) | <0.001 | 2455 |
|  | DII | Model 2: per 1-SD higher score | -0.134 (-0.177 to -0.090) | <0.001 | 2455 |
|  | DII | Model 3: per 1-SD higher score | -0.091 (-0.143 to -0.040) | 0.005 | 2455 |
| B. Weighted quartiles and global cognitive performance | FISI | Q2 vs Q1 | -0.036 (-0.230 to 0.157) | 0.630 | 2455 |
|  | FISI | Q3 vs Q1 | -0.044 (-0.231 to 0.143) | 0.549 | 2455 |
|  | FISI | Q4 vs Q1 | -0.031 (-0.209 to 0.146) | 0.649 | 2455 |
|  | FISI | P for trend |  | 0.585 | 2455 |
|  | DII | Q2 vs Q1 | -0.057 (-0.254 to 0.141) | 0.470 | 2455 |
|  | DII | Q3 vs Q1 | -0.179 (-0.340 to -0.018) | 0.037 | 2455 |
|  | DII | Q4 vs Q1 | -0.219 (-0.383 to -0.056) | 0.020 | 2455 |
|  | DII | P for trend |  | 0.007 | 2455 |
| C. Low cognitive performance | FISI | Model 3: per 1-SD higher score | 0.987 (0.846 to 1.152) | 0.846 | 2455 |
|  | DII | Model 3: per 1-SD higher score | 1.111 (0.936 to 1.319) | 0.184 | 2455 |

Continuous-outcome estimates are regression coefficients ( $\beta$ ) for differences in global cognitive Z-score; binary-outcome estimates are odds ratios (ORs) for low cognitive performance. Model 1 adjusted for age and sex. Model 2 additionally adjusted for race/ethnicity, education, marital status, PIR, and PIR missingness. Model 3 additionally adjusted for smoking, alcohol intake and missingness, recreational physical activity, BMI, two-day mean energy intake, hypertension, diabetes, and hyperlipidemia. Exposures were standardized using survey-weighted means and SDs. BMI, body mass index; CI, confidence interval; DII, Dietary Inflammatory Index; FISI, Food Inflammation Score for Individuals; PIR, poverty-income ratio; SD, standard deviation.

For DII, the corresponding estimates were −0.230 (95% CI, −0.275 to −0.185; P < 0.001) in Model 1, −0.134 (95% CI, −0.177 to −0.090; P < 0.001) in Model 2, and −0.091 (95% CI, −0.143 to −0.040; P = 0.005) in Model 3. In the fully adjusted quartile analysis, FISI showed no monotonic pattern (P for trend = 0.585). For DII, the Q3 and Q4 differences versus Q1 were −0.179 (95% CI, −0.340 to −0.018; P = 0.037) and −0.219 (95% CI, −0.383 to −0.056; P = 0.020), respectively, with P for trend = 0.007.

### 3.3 Cognitive domains, mutual adjustment, and dose-response

FISI was not associated with any domain in fully adjusted models (Figure 2; Supplementary Table S3). Higher DII was nominally associated with lower CERAD immediate recall (β = −0.062; P = 0.041), CERAD delayed recall (β = −0.080; P = 0.026), and Animal Fluency (β = −0.092; P = 0.033), but none of the four domain results remained significant at FDR <0.05. The DSST estimate was −0.054 (P = 0.094).

**Figure 2.**
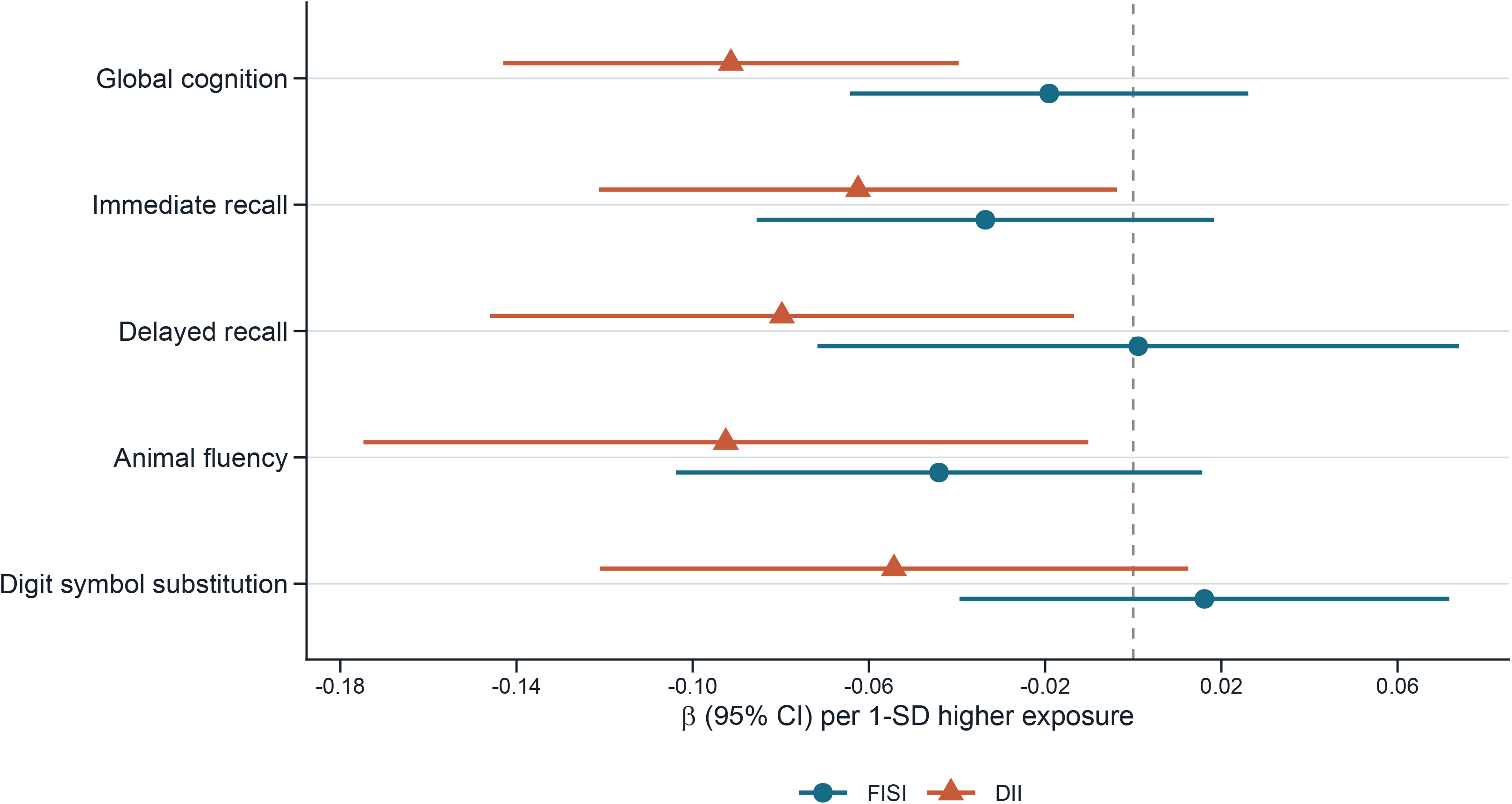
Fully adjusted associations of FISI and DII with global and domain-specific cognitive performance. Points are survey-weighted regression coefficients and horizontal bars are 95% confidence intervals per 1-SD higher exposure. Models adjusted for age, sex, race/ethnicity, education, marital status, poverty-income ratio and missingness, smoking, alcohol intake and missingness, recreational physical activity, body mass index, two-day mean energy intake, hypertension, diabetes, and hyperlipidemia. Domain-specific nominal P values were corrected across four domains within each exposure using the Benjamini-Hochberg false discovery rate; none remained significant at 0.05. Preferred final width: two columns.

When both scores were entered into Model 3, the FISI estimate was 0.021 (95% CI, −0.038 to 0.079; P = 0.403), whereas the DII estimate was −0.103 (95% CI, −0.173 to −0.032; P = 0.014) (Supplementary Table S4). Restricted cubic splines did not support non-linearity for either score (P for non-linearity = 0.834 for FISI and 0.853 for DII; Supplementary Figure S3).

### 3.4 Low cognitive performance and discrimination

In fully adjusted logistic models, neither FISI (OR per SD, 0.987; 95% CI, 0.846-1.152; P = 0.846) nor DII (OR per SD, 1.111; 95% CI, 0.936-1.319; P = 0.184) was associated with low cognitive performance (Table 2).

The survey-weighted AUROC was 0.8242 (95% bootstrap CI, 0.8110-0.8469) for the base model, 0.8242 (95% bootstrap CI, 0.8111-0.8471) after adding FISI, and 0.8247 (95% bootstrap CI, 0.8110-0.8481) after adding DII (Table 3; Supplementary Figure S1). The AUROC difference was −0.00002 for FISI versus the base model (95% bootstrap CI, −0.00020 to 0.00074; P = 0.656) and 0.00051 for DII versus the base model (95% bootstrap CI, −0.00029 to 0.00213; P = 0.280). The difference between FISI- and DII-augmented models was −0.00053 (95% bootstrap CI, −0.00213 to 0.00056; P = 0.448). Calibration patterns were also similar (Supplementary Figure S4).

**Table 3.** Survey-weighted discrimination for low cognitive performance.

| Comparison | Metric | Estimate (95% bootstrap CI) | Bootstrap P |
| --- | --- | --- | --- |
| Base | Survey-weighted AUROC | 0.8242 (0.8110 to 0.8469) |  |
| Base + FISI | Survey-weighted AUROC | 0.8242 (0.8111 to 0.8471) |  |
| Base + DII | Survey-weighted AUROC | 0.8247 (0.8110 to 0.8481) |  |
| FISI vs Base | Difference in survey-weighted AUROC | -0.0000 (-0.0002 to 0.0007) | 0.656 |
| DII vs Base | Difference in survey-weighted AUROC | 0.0005 (-0.0003 to 0.0021) | 0.280 |
| FISI vs DII | Difference in survey-weighted AUROC | -0.0005 (-0.0021 to 0.0006) | 0.448 |

*The base model included all Model 3 covariates. Confidence intervals and two-sided P values used 500 replicates that resampled primary sampling units with replacement within strata and refitted each model. AUROC differences are not equivalence tests. AUROC, area under the receiver operating characteristic curve; CI, confidence interval; DII, Dietary Inflammatory Index; FISI, Food Inflammation Score for Individuals; NA, not applicable.*

### 3.5 Subgroup and sensitivity analyses

There was no interaction of FISI or DII with diabetes or NHANES cycle, and no FISI interaction with obesity (all nominal P ≥0.195). The nominal DII-by-obesity interaction had P = 0.043 but did not remain significant after FDR correction (P_FDR = 0.260); it was therefore not interpreted as established heterogeneity (Supplementary Tables S6 and S7).

The overall pattern was stable in sensitivity analyses (Supplementary Table S5; Supplementary Figure S2). In complete covariate cases (n = 1,281), FISI remained null (β = −0.007; P = 0.683), whereas the DII association was stronger (β = −0.133; 95% CI, −0.196 to −0.070; P < 0.001). DII remained inversely associated with global cognition after excluding participants with self-reported stroke or coronary heart disease (β = −0.096; P = 0.011) and among participants with flavonoid gram coverage ≥95% (β = −0.087; P = 0.011). Cycle-specific estimates differed in nominal significance, but formal cross-cycle interactions were not significant. Alternative thresholds for low cognitive performance did not materially change the binary-outcome conclusion (Supplementary Table S8).

## 4. Discussion

### 4.1 Principal findings

In this nationally representative cross-sectional analysis of older adults, the food-based FISI and nutrient-based DII produced different fully adjusted association patterns. Higher DII was associated with lower global cognitive performance, with a significant quartile trend, whereas the FISI association attenuated after socioeconomic and behavioral adjustment. Neither score was associated with the binary low-performance threshold after full adjustment, and neither materially improved survey-weighted discrimination beyond established covariates. These results do not support treating FISI and DII as equivalent or interchangeable.

### 4.2 Comparison with previous evidence

The DII result is directionally consistent with earlier NHANES analyses that linked pro-inflammatory diets to poorer performance on selected cognitive tests [7–9]. It also accords with longitudinal observations from the HELIAD and UK Biobank cohorts, in which higher inflammatory dietary scores were associated with incident dementia [10,11], and with meta-analytic evidence suggesting adverse cognitive outcomes among people with more pro-inflammatory diets [3,15]. The present estimate was modest—approximately 0.09 SD lower global cognition per 1-SD higher DII—and domain-specific associations did not survive FDR correction. This magnitude and multiplicity pattern argue for a cautious epidemiological interpretation rather than a diagnostic or therapeutic claim.

### 4.3 Why FISI and DII may diverge

The null fully adjusted FISI finding is informative because FISI and DII were calculated from the same two dietary days and analyzed in the same participants, with identical survey weights and covariates. Their weighted correlation of 0.726 indicates substantial shared ranking, but correlation alone does not establish construct identity. DII standardizes individual two-day parameter intakes against global reference distributions before applying inflammatory weights. In contrast, FISI first assigns each food an inflammatory value per 100 g and then sums food-specific values according to grams consumed. These operations can emphasize different aspects of dietary composition and quantity. The persistence of DII, but not FISI, in the mutually adjusted model further suggests that the two measures did not contribute identical information in this dataset.

Measurement features may also have contributed to the discordance. FISI used 39 food components, including five flavonoid subclasses linked from a USDA database developed for 2007-2010 food codes. Although 89.8% of consumed grams were covered and a ≥95% coverage sensitivity analysis did not reveal a FISI association, assigning zero to unmatched flavonoids may have introduced non-differential exposure error. Food-level winsorization reduced the influence of extreme composition values, but it also represents an analytic choice; using un-winsorized values did not materially alter the conclusion. DII had a different limitation: only 33 of its 45 candidate parameters were available. Thus, neither implementation should be viewed as a complete or error-free measure of inflammatory dietary potential.

### 4.4 Outcome definition and discrimination

Our continuous and binary results also illustrate the consequences of outcome definition. A standardized global score retains information across the cognitive distribution and combined four tests, whereas the weighted 25th-percentile threshold discards gradation and does not establish a clinical diagnosis. Earlier NHANES reports used different test-specific thresholds, component counts, samples, or adjustment sets [7–9], which may partly explain why their binary findings were not reproduced in our fully adjusted global-threshold model. The lack of incremental AUROC change provides a separate message: even the DII association did not translate into measurable discrimination beyond a strong base model. AUROC is not the sole criterion for evaluating a risk factor [25], but the observed changes and bootstrap intervals offer no basis for presenting either score as a clinical prediction tool.

### 4.5 Strengths and limitations

This study has several strengths. Both exposures used two reliable recalls and a two-day energy covariate, the analysis respected the combined-cycle dietary weights and complex sampling design, and the same frozen sample and covariate hierarchy were applied to both scores. The FISI calculation was audited component by component and independently reconstructed food by food for ten participants. We reported the actual 33 DII parameters, quantified flavonoid coverage, corrected domain and interaction families for multiple testing, and used stratified primary-sampling-unit bootstrap refitting for discrimination.

Several limitations remain. First, the cross-sectional design prevents temporal or causal inference; poorer cognition could influence food choice or recall quality. Second, two 24-h recalls do not characterize long-term usual intake and are vulnerable to within-person variation and reporting error. Third, the food composition and flavonoid linkage may not fully reflect all foods consumed in 2011-2014, and FISI has limited independent epidemiological validation. Fourth, using 33 DII parameters may limit comparability with studies using different parameter sets. Fifth, imputation or unknown categories retained participants but cannot eliminate bias from missing covariates, although complete-case results were consistent. Sixth, residual confounding by unmeasured health, medication, social, or dietary factors is possible. Finally, the cognitive battery and percentile threshold measure performance rather than a clinical disorder, and exclusions required for complete dietary and cognitive data may limit generalizability.

## 5. Conclusion

Higher DII was associated with lower global cognitive performance after comprehensive adjustment, whereas FISI was not. Neither score was associated with fully adjusted low cognitive performance or materially improved discrimination beyond established covariates. Food- and nutrient-based inflammatory scores captured overlapping but non-interchangeable information in this cross-sectional sample. Prospective studies with repeated dietary assessment and updated food-composition linkage are needed before either measure is considered for cognitive risk stratification.

## Declarations

### Ethics approval and consent to participate

The NCHS Ethics Review Board approved NHANES 2011-2012 under Protocol #2011-17 and NHANES 2013-2014 as a continuation of that protocol. All participants provided written informed consent. This secondary analysis used deidentified public-use data.

### Consent for publication

Not applicable.

## Author contributions

- **Jian Zheng:** Conceptualization; Methodology; Software; Formal analysis; Visualization; Writing – original draft.
- **Jiayu Du:** Validation; Investigation; Writing – review & editing.
- **Zijie Liu:** Data curation; Validation; Writing – review & editing.
- **Shuyue Jin:** Investigation; Resources; Writing – review & editing.
- **Kaiguo Tian:** Methodology; Supervision; Writing – review & editing.
- **Jingxuan Wang:** Validation; Project administration; Writing – review & editing.
- **Qi Zhang:** Conceptualization; Supervision; Funding acquisition; Project administration; Writing – review & editing.

## Declaration of generative AI and AI-assisted technologies

During the preparation of this work, the authors used ChatGPT and Anthropic Claude solely to polish the language. After using these tools, the authors reviewed and edited the content as needed and take full responsibility for the content of the published article.

## Data Availability

The NHANES public-use data and documentation are available from the National Center for Health Statistics (https://www.cdc.gov/nchs/nhanes/). The analysis code and non-identifying derived materials are available from the corresponding author on reasonable request.

