## Supplementary Material for "Discordant Associations of Food- and Nutrient-Based Dietary Inflammatory Scores With Cognitive Performance in Older Adults: A Cross-Sectional Study"

### Supplementary methods

#### Food-composition linkage and FISI audit

The FII calculation used 39 food components: 34 nutrient variables from the cycle-specific NHANES individual-food files and five flavonoid subclasses from the USDA Database of Flavonoid Values for USDA Food Codes 2007-2010. Component values were expressed per 100 g. For a unique food code appearing more than once within a cycle, the median per-100-g component value was used. Raw food-level FII values were winsorized at the cycle-specific 1st and 99th percentiles for the primary analysis.

Flavonoid values were linked by eight-digit USDA food code. Unmatched values were set to zero. Coverage was quantified both as the proportion of food records linked and as the proportion of consumed grams linked. A sensitivity analysis retained participants with ≥95% gram-weighted coverage.

Ten participants spanning the observed FISI distribution were selected for independent verification. For each person and day, the verification loop added each food's winsorized FII multiplied by grams consumed divided by 100. Day 1 and Day 2 sums were then averaged. This second implementation was compared with the production vectorized calculation.

#### DII implementation audit

Thirty-three of the 45 candidate DII parameters were present in both cycles and dietary days. Daily source values came from the NHANES total nutrient files, except flavonoids, which were calculated from individual-food records. Participant values were two-day means. All 33 parameters were standardized against the published global means and standard deviations, converted to centerd percentiles, multiplied by their inflammatory effect scores, and summed. Every participant in the final analytic set had all 33 implemented parameters.

### Sensitivity and subgroup inference

Sensitivity models repeated the fully adjusted continuous-outcome analysis in complete covariate cases; after excluding self-reported stroke or coronary heart disease; among participants with flavonoid gram coverage ≥95%; without food-level winsorization; using energy-scaled FISI per 1,000 kcal; and within each NHANES cycle. The separate-cycle models used the original two-year WTDR2D weight. Because the fully adjusted single-cycle models exhausted the survey residual degrees of freedom, their confidence intervals and P values used normal-approximation Wald inference; cross-cycle heterogeneity was evaluated in the combined design.

Diabetes and obesity interactions omitted the corresponding defining covariate from the adjustment set. Cycle interaction models used the full covariate set. Six interaction P values were corrected with the Benjamini-Hochberg false discovery rate. Within-stratum significance was not used as evidence of heterogeneity.

### Discrimination analysis

The base survey-weighted quasibinomial model included age, sex, race/ethnicity, education, marital status, poverty-income ratio and missingness, smoking, alcohol intake and missingness, recreational physical activity, body mass index, two-day mean energy, hypertension, diabetes, and hyperlipidemia. FISI or DII was then added separately. Survey-weighted AUROCs were calculated from model predictions. For inference, primary sampling units were sampled with replacement within each stratum, their weights were multiplied by sampling multiplicity, each model was refitted, and AUROCs were recalculated in 500 replicates.

### Table S1. Sequential participant selection

| Step | Participants remaining | Excluded from previous step |
|---|---|---|
| NHANES 2011-2014 participants | 19931 | |
| Age ≥60 years | 3632 | 16299 |
| All four cognitive tests complete | 2934 | 698 |
| Two reliable recalls (DRDINT=2; both status=1) | 2524 | 410 |
| WTDR2D>0 and survey design variables complete | 2524 | 0 |
| Two-day FISI and 33-parameter DII calculable | 2524 | 0 |
| Plausible sex-specific two-day mean energy | 2508 | 16 |
| BMI 15-50 kg/m² and core demographics complete | 2455 | 53 |

*Counts are unweighted. Exclusions were applied sequentially to NHANES 2011-2014 participants aged 60 years or older. BMI, body mass index; DII, Dietary Inflammatory Index; FISI, Food Inflammation Score for Individuals; NHANES, National Health and Nutrition Examination Survey.*

### Table S2. Exposure-construction and method audit

| Check | Value |
|---|---|
| G0 version | Frozen v3.0 (2026-07-30) |
| Final unweighted N | 2455 |
| All DRDINT=2 | TRUE |
| All DR1DRSTZ=1 | TRUE |
| All DR2DRSTZ=1 | TRUE |
| All WTDR2D>0 | TRUE |
| All n_diet_days=2 | TRUE |
| Weight formula | WTDR2D/2 |
| Energy window | Mean of Day 1 and Day 2 kcal |
| FISI formula | mean_day[sum_food(FII_per100g*grams/100)] |
| Individual FISI normalized to 1-100 | FALSE |
| Individual FISI divided by total grams | FALSE |
| FISI has negative values | TRUE |
| DII available parameters | 33 |
| FII food components | 39 |
| Physical activity coding | active/inactive/unknown (no MET-min/week) |
| Low cognitive performance threshold | Survey-weighted 25th percentile of Global Z = -0.697375 |

| Check | Value |
|---|---|
| FISI verification participants | 10 |
| Maximum verification difference | 1.776357e-15 |
| PIR imputed N | 186 |
| Alcohol imputed N | 1063 |
| Flavonoid coverage: 2011-2012 | record 90.7%; gram-weighted 93.3%; 5191 unique food codes |
| Flavonoid coverage: 2013-2014 | record 85.9%; gram-weighted 86.3%; 5531 unique food codes |
| Flavonoid coverage: Overall | record 88.3%; gram-weighted 89.8%; 6466 unique food codes |

*This audit records the frozen v3 exposure construction and modeling decisions. DII, Dietary Inflammatory Index; FISI, Food Inflammation Score for Individuals; NHANES, National Health and Nutrition Examination Survey.*

### Table S3. Fully adjusted associations with cognitive domains

| Domain | Exposure | β (95% CI) | Nominal P value | BH-FDR P value | n |
|---|---|---|---|---|---|
| CERAD immediate recall | FISI | -0.034 (-0.085 to 0.018) | 0.165 | 0.330 | 2455 |
| CERAD delayed recall | FISI | 0.001 (-0.072 to 0.074) | 0.971 | 0.971 | 2455 |
| Animal fluency | FISI | -0.044 (-0.104 to 0.016) | 0.121 | 0.330 | 2455 |
| DSST | FISI | 0.016 (-0.039 to 0.072) | 0.504 | 0.672 | 2455 |
| CERAD immediate recall | DII | -0.062 (-0.121 to -0.004) | 0.041 | 0.054 | 2455 |
| CERAD delayed recall | DII | -0.080 (-0.146 to -0.013) | 0.026 | 0.054 | 2455 |
| Animal fluency | DII | -0.092 (-0.175 to -0.010) | 0.033 | 0.054 | 2455 |
| DSST | DII | -0.054 (-0.121 to 0.012) | 0.094 | 0.094 | 2455 |

*Estimates are differences in standardized domain scores per 1-SD higher exposure. BH-FDR P values correct the four domains within each exposure. BH-FDR, Benjamini-Hochberg false discovery rate; CI, confidence interval; DII, Dietary Inflammatory Index; FISI, Food Inflammation Score for Individuals; SD, standard deviation.*

### Table S4. Mutual-adjustment and restricted-cubic-spline results

| Analysis | Exposure | β (95% CI) | P value | n |
|---|---|---|---|---|
| Mutually adjusted Model 3 | FISI | 0.021 (-0.038 to 0.079) | 0.403 | 2455 |
| Mutually adjusted Model 3 | DII | -0.103 (-0.173 to -0.032) | 0.014 | 2455 |
| Restricted cubic spline Model 3 | FISI | | overall 0.706; nonlinear 0.834 | 2455 |
| Restricted cubic spline Model 3 | DII | | overall 0.053; nonlinear 0.853 | 2455 |

*The mutual model included standardized FISI and DII together with all Model 3 covariates. Splines used survey-weighted knots at the 5th, 35th, 65th, and 95th percentiles. CI, confidence interval; DII, Dietary Inflammatory Index; FISI, Food Inflammation Score for Individuals.*

### Table S5. Continuous-outcome sensitivity analyses

| Analysis | Exposure | β (95% CI) | P value | n | Weight | Inference |
|---|---|---|---|---|---|---|
| Primary frozen sample | FISI | -0.019 (-0.064 to 0.026) | 0.341 | 2455 | WTDR2D/2 | Survey-design t inference |
| Primary frozen sample | DII | -0.091 (-0.143 to -0.040) | 0.005 | 2455 | WTDR2D/2 | Survey-design t inference |
| Complete covariate cases | FISI | -0.007 (-0.043 to 0.029) | 0.683 | 1281 | WTDR2D/2 | Survey-design t inference |
| Complete covariate cases | DII | -0.133 (-0.196 to -0.070) | <0.001 | 1281 | WTDR2D/2 | Survey-design t inference |
| FISI per 1000 kcal | FISI | -0.018 (-0.069 to 0.033) | 0.414 | 2455 | WTDR2D/2 | Survey-design t inference |
| Exclude self-reported stroke/CHD | FISI | -0.013 (-0.056 to 0.031) | 0.504 | 2096 | WTDR2D/2 | Survey-design t inference |
| Exclude self-reported stroke/CHD | DII | -0.096 (-0.161 to -0.031) | 0.011 | 2096 | WTDR2D/2 | Survey-design t inference |
| Flavonoid gram coverage ≥95% | FISI | -0.033 (-0.101 to 0.035) | 0.299 | 1234 | WTDR2D/2 | Survey-design t inference |
| Flavonoid gram coverage ≥95% | DII | -0.087 (-0.148 to -0.025) | 0.011 | 1234 | WTDR2D/2 | Survey-design t inference |
| No food-level 1st-99th percentile winsorization | FISI | -0.031 (-0.080 to 0.017) | 0.163 | 2455 | WTDR2D/2 | Survey-design t inference |
| No food-level 1st-99th percentile winsorization | DII | -0.091 (-0.143 to -0.040) | 0.005 | 2455 | WTDR2D/2 | Survey-design t inference |
| NHANES 2011-2012 | FISI | -0.018 (-0.061 to 0.025) | 0.411 | 1158 | WTDR2D | Normal-approximation Wald inference because the fully adjusted single-cycle model exhausts survey residual degrees of freedom |
| NHANES 2011-2012 | DII | -0.104 (-0.147 to -0.061) | <0.001 | 1158 | WTDR2D | Normal-approximation Wald inference because the fully adjusted single-cycle model exhausts survey residual degrees of freedom |
| NHANES 2013-2014 | FISI | -0.064 (-0.114 to -0.014) | 0.012 | 1297 | WTDR2D | Normal-approximation Wald inference because the fully adjusted single-cycle model exhausts survey residual degrees of freedom |

| Analysis | Exposure | β (95% CI) | P value | n | Weight | Inference |
|---|---|---|---|---|---|---|
| NHANES 2013-2014 | DII | -0.108 (-0.181 to -0.035) | 0.004 | 1297 | WTDR2D | Normal-approximation Wald inference because the fully adjusted single-cycle model exhausts survey residual degrees of freedom |

*Estimates are differences in global cognitive Z-score per 1-SD higher exposure. Separate-cycle rows used WTDR2D and normal-approximation Wald inference because fully adjusted single-cycle models exhausted survey residual degrees of freedom. CHD, coronary heart disease; CI, confidence interval; DII, Dietary Inflammatory Index; FISI, Food Inflammation Score for Individuals; NHANES, National Health and Nutrition Examination Survey; SD, standard deviation.*

### Table S6. Subgroup interaction tests

| Subgroup | Exposure | Interaction β (95% CI) | Interaction P value | BH-FDR P value | n |
|---|---|---|---|---|---|
| Diabetes | FISI | 0.053 (-0.079 to 0.184) | 0.349 | 0.422 | 2455 |
| Diabetes | DII | 0.057 (-0.055 to 0.168) | 0.247 | 0.422 | 2455 |
| Obesity | FISI | 0.045 (-0.075 to 0.165) | 0.379 | 0.422 | 2455 |
| Obesity | DII | 0.112 (0.005 to 0.220) | 0.043 | 0.260 | 2455 |
| NHANES cycle | FISI | -0.069 (-0.193 to 0.054) | 0.195 | 0.422 | 2455 |
| NHANES cycle | DII | -0.031 (-0.128 to 0.066) | 0.422 | 0.422 | 2455 |

*Interaction coefficients are differences in exposure slopes between the displayed subgroup levels. BH-FDR P values correct the six interaction tests. Within-stratum significance was not used as evidence of heterogeneity. BH-FDR, Benjamini-Hochberg false discovery rate; CI, confidence interval; DII, Dietary Inflammatory Index; FISI, Food Inflammation Score for Individuals.*

### Table S7. Subgroup stratum-specific estimates

| Subgroup | Exposure | β (95% CI) | P value | n |
|---|---|---|---|---|
| Diabetes=0 | FISI | -0.026 (-0.072 to 0.021) | 0.237 | 1780 |
| Diabetes=1 | FISI | 0.055 (-0.063 to 0.172) | 0.315 | 675 |
| Diabetes=0 | DII | -0.116 (-0.179 to -0.053) | 0.003 | 1780 |
| Diabetes=1 | DII | 0.011 (-0.093 to 0.116) | 0.809 | 675 |
| Obesity=0 | FISI | -0.019 (-0.060 to 0.021) | 0.295 | 1518 |
| Obesity=1 | FISI | 0.002 (-0.102 to 0.107) | 0.964 | 937 |
| Obesity=0 | DII | -0.126 (-0.208 to -0.043) | 0.008 | 1518 |
| Obesity=1 | DII | -0.029 (-0.110 to 0.052) | 0.432 | 937 |

*Estimates are differences in global cognitive Z-score per 1-SD higher exposure within the displayed strata. CI, confidence interval; DII, Dietary Inflammatory Index; FISI, Food Inflammation Score for Individuals; SD, standard deviation.*

### Table S8. Alternative thresholds for low cognitive performance

| Threshold | Global Z cut-off | Exposure | OR (95% CI) | P value | n |
| --- | --- | --- | --- | --- | --- |
| Weighted 20th percentile | -0.857 | FISI | 1.070 (0.892 to 1.283) | 0.398 | 2455 |
| Weighted 20th percentile | -0.857 | DII | 1.176 (0.990 to 1.397) | 0.061 | 2455 |
| Weighted 30th percentile | -0.497 | FISI | 1.024 (0.880 to 1.192) | 0.715 | 2455 |
| Weighted 30th percentile | -0.497 | DII | 1.130 (0.945 to 1.351) | 0.146 | 2455 |

*Odds ratios compare the odds of low cognitive performance per 1-SD higher exposure using the displayed alternative outcome thresholds and the fully adjusted covariate set. CI, confidence interval; DII, Dietary Inflammatory Index; FISI, Food Inflammation Score for Individuals; OR, odds ratio; SD, standard deviation.*

### Table S9. Thirty-three DII parameters used in the v3 analysis

| Parameter | Key | Global mean | Global SD | Inflammatory effect score | Nonmissing source records |
|---|---|---|---|---|---|
| Alcohol (g) | ALCO | 13.980 | 3.720 | -0.278 | 14935 |
| Vitamin B12 (μg) | VB12 | 5.150 | 2.700 | 0.106 | 14935 |
| Vitamin B6 (mg) | VB6 | 1.470 | 0.740 | -0.365 | 14935 |
| β-Carotene (μg) | BCAR | 3718.000 | 1720.000 | -0.584 | 14935 |
| Caffeine (g) | CAFF | 8.050 | 6.670 | -0.110 | 14935 |
| Carbohydrate (g) | CARB | 272.200 | 40.000 | 0.097 | 14935 |
| Cholesterol (mg) | CHOL | 279.400 | 51.200 | 0.110 | 14935 |
| Energy (kcal) | KCAL | 2056.000 | 338.000 | 0.180 | 14935 |
| Total fat (g) | TFAT | 71.400 | 19.400 | 0.298 | 14935 |
| Fiber (g) | FIBE | 18.800 | 4.900 | -0.663 | 14935 |
| Folic acid (μg) | FA | 273.000 | 70.700 | -0.190 | 14935 |
| Fe (mg) | IRON | 13.350 | 3.710 | 0.032 | 14935 |
| Mg (mg) | MAGN | 310.100 | 139.400 | -0.484 | 14935 |
| MUFA (g) | MFAT | 27.000 | 6.100 | -0.009 | 14935 |
| Niacin (mg) | NIAC | 25.900 | 11.770 | -0.246 | 14935 |
| n-3 Fatty acids (g) | N3 | 1.060 | 1.060 | -0.436 | 14935 |
| n-6 Fatty acids (g) | N6 | 10.800 | 7.500 | -0.159 | 14935 |

| Parameter | Key | Global mean | Global SD | Inflammatory effect score | Nonmissing source records |
|---|---|---|---|---|---|
| Protein (g) | PROT | 79.400 | 13.900 | 0.021 | 14935 |
| PUFA (g) | PFAT | 13.880 | 3.760 | -0.337 | 14935 |
| Riboflavin (mg) | VB2 | 1.700 | 0.790 | -0.068 | 14935 |
| Saturated fat (g) | SFAT | 28.600 | 8.000 | 0.373 | 14935 |
| Se (μg) | SELE | 67.000 | 25.100 | -0.191 | 14935 |
| Thiamin (mg) | VB1 | 1.700 | 0.660 | -0.098 | 14935 |
| Vitamin A (RE) | VARA | 983.900 | 518.600 | -0.401 | 14935 |
| Vitamin C (mg) | VC | 118.200 | 43.460 | -0.424 | 14935 |
| Vitamin D (μg) | VD | 6.260 | 2.210 | -0.446 | 14935 |
| Vitamin E (mg) | ATOC | 8.730 | 1.490 | -0.419 | 14935 |
| Zn (mg) | ZINC | 9.840 | 2.190 | -0.313 | 14935 |
| Flavan-3-ol (mg) | FLAV_Flavan-3-ols | 95.800 | 85.900 | -0.415 | 15106 |
| Flavones (mg) | FLAV_Flavones | 1.550 | 0.070 | -0.616 | 15106 |
| Flavonols (mg) | FLAV_Flavonols | 17.700 | 6.790 | -0.467 | 15106 |
| Flavonones (mg) | FLAV_Flavanones | 11.700 | 3.820 | -0.250 | 15106 |
| Isoflavones (mg) | FLAV_Isoflavones | 1.200 | 0.200 | -0.593 | 15106 |

*Global reference means, standard deviations, and inflammatory effect scores are the fixed DII parameters used to construct the v3 exposure. DII, Dietary Inflammatory Index; SD, standard deviation.*

### Table S10. Independent food-by-food FISI verification in ten participants

#### Panel A. Day-specific verification

| Masked participant ID | Program Day 1 | Independent Day 1 | Absolute difference Day 1 | Program Day 2 | Independent Day 2 | Absolute difference Day 2 | Food rows |
|---|---|---|---|---|---|---|---|
| V01 | -111.617413 | -111.617413 | 0.00e+00 | -157.969850 | -157.969850 | 2.84e-14 | 36 |
| V02 | -7.742833 | -7.742833 | 8.88e-16 | -23.362033 | -23.362033 | 0.00e+00 | 36 |
| V03 | -12.684849 | -12.684849 | 0.00e+00 | -11.798914 | -11.798914 | 1.78e-15 | 37 |
| V04 | -6.351707 | -6.351707 | 8.88e-16 | -13.829126 | -13.829126 | 0.00e+00 | 55 |
| V05 | -8.500985 | -8.500985 | 0.00e+00 | -8.794453 | -8.794453 | 1.78e-15 | 52 |
| V06 | -9.653233 | -9.653233 | 1.78e-15 | -5.343368 | -5.343368 | 0.00e+00 | 46 |
| V07 | -7.462415 | -7.462415 | 8.88e-16 | -5.424621 | -5.424621 | 8.88e-16 | 29 |
| V08 | -6.414746 | -6.414746 | 8.88e-16 | -4.439231 | -4.439231 | 8.88e-16 | 21 |
| V09 | -3.685715 | -3.685715 | 0.00e+00 | -4.478908 | -4.478908 | 0.00e+00 | 23 |
| V10 | -0.558072 | -0.558072 | 1.11e-16 | -1.090390 | -1.090390 | 0.00e+00 | 18 |

#### Panel B. Two-day mean verification

| Masked participant ID | Program two-day mean | Independent two-day mean | Absolute difference two-day mean |
|---|---|---|---|
| V01 | -134.793631 | -134.793631 | 0.00e+00 |
| V02 | -15.552433 | -15.552433 | 1.78e-15 |
| V03 | -12.241882 | -12.241882 | 0.00e+00 |

| Masked participant ID | Program two-day mean | Independent two-day mean | Absolute difference two-day mean |
|---|---|---|---|
| V04 | -10.090416 | -10.090416 | 0.00e+00 |
| V05 | -8.647719 | -8.647719 | 0.00e+00 |
| V06 | -7.498301 | -7.498301 | 8.88e-16 |
| V07 | -6.443518 | -6.443518 | 1.78e-15 |
| V08 | -5.426988 | -5.426988 | 0.00e+00 |
| V09 | -4.082311 | -4.082311 | 0.00e+00 |
| V10 | -0.824231 | -0.824231 | 0.00e+00 |

*Masked verification identifiers replace NHANES sequence numbers. Program and independent values are displayed to six decimal places; absolute differences use scientific notation. FISI, Food Inflammation Score for Individuals; NHANES, National Health and Nutrition Examination Survey.*

### Table S11. FII component mapping - NHANES 2011-2012

| Cycle | Component | Key | Source variable | Nonmissing, n | Nonzero, n |
|---|---|---|---|---|---|
| 2011-2012 | Protein | PROT | NHANES IFF | 5191 | 5081 |
| 2011-2012 | Total lipid (fat) | TFAT | NHANES IFF | 5191 | 5020 |
| 2011-2012 | Carbohydrate, by difference | CARB | NHANES IFF | 5191 | 4905 |
| 2011-2012 | Energy | KCAL | NHANES IFF | 5191 | 5174 |
| 2011-2012 | Alcohol, ethyl | ALCO | NHANES IFF | 5191 | 69 |
| 2011-2012 | Caffeine | CAFF | NHANES IFF | 5191 | 366 |
| 2011-2012 | Fiber, total dietary | FIBE | NHANES IFF | 5191 | 4021 |
| 2011-2012 | Iron, Fe | IRON | NHANES IFF | 5191 | 5074 |
| 2011-2012 | Magnesium, Mg | MAGN | NHANES IFF | 5191 | 5017 |
| 2011-2012 | Zinc, Zn | ZINC | NHANES IFF | 5191 | 5068 |
| 2011-2012 | Selenium, Se | SELE | NHANES IFF | 5191 | 4948 |
| 2011-2012 | Vitamin A, RAE | VARA | NHANES IFF | 5191 | 4012 |
| 2011-2012 | Carotene, alpha | ACAR | NHANES IFF | 5191 | 1485 |
| 2011-2012 | Vitamin E (alpha-tocopherol) | ATOC | NHANES IFF | 5191 | 4896 |
| 2011-2012 | Vitamin D (D2 + D3) | VD | NHANES IFF | 5191 | 2455 |
| 2011-2012 | Vitamin C, total ascorbic acid | VC | NHANES IFF | 5191 | 3515 |
| 2011-2012 | Thiamin | VB1 | NHANES IFF | 5191 | 4982 |
| 2011-2012 | Riboflavin | VB2 | NHANES IFF | 5191 | 5048 |

| Cycle | Component | Key | Source variable | Nonmissing, n | Nonzero, n |
|---|---|---|---|---|---|
| 2011-2012 | Niacin | NIAC | NHANES IFF | 5191 | 5046 |
| 2011-2012 | Vitamin B-6 | VB6 | NHANES IFF | 5191 | 4969 |
| 2011-2012 | Vitamin B-12 | VB12 | NHANES IFF | 5191 | 3426 |
| 2011-2012 | Folic acid | FA | NHANES IFF | 5191 | 1933 |
| 2011-2012 | Vitamin E, added | ATOA | NHANES IFF | 5191 | 173 |
| 2011-2012 | Vitamin B-12, added | B12A | NHANES IFF | 5191 | 330 |
| 2011-2012 | Cholesterol | CHOL | NHANES IFF | 5191 | 2924 |
| 2011-2012 | Fatty acids, total saturated | SFAT | NHANES IFF | 5191 | 4971 |
| 2011-2012 | PUFA 18:2 | P182 | NHANES IFF | 5191 | 4974 |
| 2011-2012 | PUFA 18:3 | P183 | NHANES IFF | 5191 | 4783 |
| 2011-2012 | PUFA 20:4 | P204 | NHANES IFF | 5191 | 2244 |
| 2011-2012 | PUFA 22:6 n-3 (DHA) | P226 | NHANES IFF | 5191 | 1473 |
| 2011-2012 | PUFA 20:5 n-3 (EPA) | P205 | NHANES IFF | 5191 | 1220 |
| 2011-2012 | PUFA 22:5 n-3 (DPA) | P225 | NHANES IFF | 5191 | 1461 |
| 2011-2012 | Fatty acids, total monounsaturated | MFAT | NHANES IFF | 5191 | 4959 |
| 2011-2012 | Fatty acids, total polyunsaturated | PFAT | NHANES IFF | 5191 | 4993 |
| 2011-2012 | Flavan-3-ols | FLAV_Flavan-3-ols | USDA flavonoid 2007-2010 | 5191 | 985 |
| 2011-2012 | Flavanones | FLAV_Flavanones | USDA flavonoid 2007-2010 | 5191 | 421 |

| Cycle | Component | Key | Source variable | Nonmissing, n | Nonzero, n |
|---|---|---|---|---|---|
| 2011-2012 | Flavones | FLAV_Flavones | USDA flavonoid 2007-2010 | 5191 | 1241 |
| 2011-2012 | Flavonols | FLAV_Flavonols | USDA flavonoid 2007-2010 | 5191 | 2386 |
| 2011-2012 | Isoflavones | FLAV_Isoflavones | USDA flavonoid 2007-2010 | 5191 | 391 |

*The table maps each NHANES cycle-specific food-code component to the source variable used in the frozen v3 FISI construction. FII, Food Inflammation Index; FISI, Food Inflammation Score for Individuals; NHANES, National Health and Nutrition Examination Survey.*

### Table S11 (continued). FII component mapping - NHANES 2013-2014

| Cycle | Component | Key | Source variable | Nonmissing, n | Nonzero, n |
|---|---|---|---|---|---|
| 2013-2014 | Protein | PROT | NHANES IFF | 5531 | 5368 |
| 2013-2014 | Total lipid (fat) | TFAT | NHANES IFF | 5531 | 5316 |
| 2013-2014 | Carbohydrate, by difference | CARB | NHANES IFF | 5531 | 5208 |
| 2013-2014 | Energy | KCAL | NHANES IFF | 5531 | 5501 |
| 2013-2014 | Alcohol, ethyl | ALCO | NHANES IFF | 5531 | 73 |
| 2013-2014 | Caffeine | CAFF | NHANES IFF | 5531 | 422 |
| 2013-2014 | Fiber, total dietary | FIBE | NHANES IFF | 5531 | 4226 |
| 2013-2014 | Iron, Fe | IRON | NHANES IFF | 5531 | 5386 |
| 2013-2014 | Magnesium, Mg | MAGN | NHANES IFF | 5531 | 5330 |
| 2013-2014 | Zinc, Zn | ZINC | NHANES IFF | 5531 | 5406 |
| 2013-2014 | Selenium, Se | SELE | NHANES IFF | 5531 | 5236 |
| 2013-2014 | Vitamin A, RAE | VARA | NHANES IFF | 5531 | 4269 |
| 2013-2014 | Carotene, alpha | ACAR | NHANES IFF | 5531 | 1569 |
| 2013-2014 | Vitamin E (alpha-tocopherol) | ATOC | NHANES IFF | 5531 | 5188 |
| 2013-2014 | Vitamin D (D2 + D3) | VD | NHANES IFF | 5531 | 2467 |
| 2013-2014 | Vitamin C, total ascorbic acid | VC | NHANES IFF | 5531 | 3751 |
| 2013-2014 | Thiamin | VB1 | NHANES IFF | 5531 | 5301 |
| 2013-2014 | Riboflavin | VB2 | NHANES IFF | 5531 | 5368 |

| Cycle | Component | Key | Source variable | Nonmissing, n | Nonzero, n |
|---|---|---|---|---|---|
| 2013-2014 | Niacin | NIAC | NHANES IFF | 5531 | 5376 |
| 2013-2014 | Vitamin B-6 | VB6 | NHANES IFF | 5531 | 5266 |
| 2013-2014 | Vitamin B-12 | VB12 | NHANES IFF | 5531 | 3413 |
| 2013-2014 | Folic acid | FA | NHANES IFF | 5531 | 1945 |
| 2013-2014 | Vitamin E, added | ATOA | NHANES IFF | 5531 | 197 |
| 2013-2014 | Vitamin B-12, added | B12A | NHANES IFF | 5531 | 326 |
| 2013-2014 | Cholesterol | CHOL | NHANES IFF | 5531 | 3238 |
| 2013-2014 | Fatty acids, total saturated | SFAT | NHANES IFF | 5531 | 5239 |
| 2013-2014 | PUFA 18:2 | P182 | NHANES IFF | 5531 | 5253 |
| 2013-2014 | PUFA 18:3 | P183 | NHANES IFF | 5531 | 5065 |
| 2013-2014 | PUFA 20:4 | P204 | NHANES IFF | 5531 | 2452 |
| 2013-2014 | PUFA 22:6 n-3 (DHA) | P226 | NHANES IFF | 5531 | 1490 |
| 2013-2014 | PUFA 20:5 n-3 (EPA) | P205 | NHANES IFF | 5531 | 1382 |
| 2013-2014 | PUFA 22:5 n-3 (DPA) | P225 | NHANES IFF | 5531 | 1652 |
| 2013-2014 | Fatty acids, total monounsaturated | MFAT | NHANES IFF | 5531 | 5232 |
| 2013-2014 | Fatty acids, total polyunsaturated | PFAT | NHANES IFF | 5531 | 5264 |
| 2013-2014 | Flavan-3-ols | FLAV_Flavan-3-ols | USDA flavonoid 2007-2010 | 5531 | 942 |
| 2013-2014 | Flavanones | FLAV_Flavanones | USDA flavonoid 2007-2010 | 5531 | 414 |

| Cycle | Component | Key | Source variable | Nonmissing, n | Nonzero, n |
| --- | --- | --- | --- | --- | --- |
| 2013-2014 | Flavones | FLAV_Flavones | USDA flavonoid 2007-2010 | 5531 | 1119 |
| 2013-2014 | Flavonols | FLAV_Flavonols | USDA flavonoid 2007-2010 | 5531 | 2194 |
| 2013-2014 | Isoflavones | FLAV_Isoflavones | USDA flavonoid 2007-2010 | 5531 | 372 |

*The table maps each NHANES cycle-specific food-code component to the source variable used in the frozen v3 FISI construction. FII, Food Inflammation Index; FISI, Food Inflammation Score for Individuals; NHANES, National Health and Nutrition Examination Survey.*

### Table S12. Missingness audit in the final analytic set

| Variable | Missing, n | Missing (%) |
|---|---|---|
| SEQN | 0 | 0.0 |
| cycle | 0 | 0.0 |
| age | 0 | 0.0 |
| sex | 0 | 0.0 |
| race | 0 | 0.0 |
| education | 0 | 0.0 |
| pir | 186 | 7.6 |
| marital | 0 | 0.0 |
| sdmvpsu | 0 | 0.0 |
| sdmvstra | 0 | 0.0 |
| bmi | 0 | 0.0 |
| smq020 | 0 | 0.0 |
| smq040 | 1215 | 49.5 |
| alq120q | 385 | 15.7 |
| alq120u | 1062 | 43.3 |
| alq130 | 1062 | 43.3 |
| alcohol_missing | 0 | 0.0 |
| frequency_year | 1062 | 43.3 |

| Variable | Missing, n | Missing (%) |
|---|---|---|
| alcohol_g_day_raw | 1063 | 43.3 |
| pa_vig | 0 | 0.0 |
| pa_mod | 0 | 0.0 |
| diabetes_self | 0 | 0.0 |
| hypertension_self | 0 | 0.0 |
| high_cholesterol | 0 | 0.0 |
| lipid_medication | 1216 | 49.5 |
| hba1c | 73 | 3.0 |
| total_cholesterol | 112 | 4.6 |
| stroke | 0 | 0.0 |
| coronary_heart_disease | 0 | 0.0 |
| sex_f | 0 | 0.0 |
| race_f | 0 | 0.0 |
| education_f | 0 | 0.0 |
| marital_f | 0 | 0.0 |
| smoke_f | 0 | 0.0 |
| physical_activity_f | 0 | 0.0 |
| diabetes | 0 | 0.0 |
| hypertension | 0 | 0.0 |

| Variable | Missing, n | Missing (%) |
|---|---|---|
| hyperlipidemia | 0 | 0.0 |

*Counts and percentages are unweighted and refer to the final analytic sample (n = 2455). Percentages are displayed to one decimal place; the underlying frozen counts are unchanged.*

### Table S12 (continued). Missingness audit in the final analytic set

| Variable | Missing, n | Missing (%) |
|---|---|---|
| cvd_history | 0 | 0.0 |
| alcohol_g_day | 0 | 0.0 |
| alcohol_miss | 0 | 0.0 |
| CERAD_IR | 0 | 0.0 |
| CERAD_DR | 0 | 0.0 |
| AFT | 0 | 0.0 |
| DSST | 0 | 0.0 |
| DR1DRSTZ | 0 | 0.0 |
| DR2DRSTZ | 0 | 0.0 |
| DRDINT | 0 | 0.0 |
| WTDR2D | 0 | 0.0 |
| wt_diet_4yr | 0 | 0.0 |
| energy_day1 | 0 | 0.0 |
| energy_day2 | 0 | 0.0 |
| energy_2day | 0 | 0.0 |
| reliable_2day | 0 | 0.0 |
| FISI | 0 | 0.0 |
| FISI_unwinsorized | 0 | 0.0 |

| Variable | Missing, n | Missing (%) |
|---|---|---|
| n_diet_days | 0 | 0.0 |
| flav_cov | 0 | 0.0 |
| total_grams_2day | 0 | 0.0 |
| DII | 0 | 0.0 |
| dii_n_comp | 0 | 0.0 |
| n_dii_days | 0 | 0.0 |
| pir_miss | 0 | 0.0 |
| pir_imp | 0 | 0.0 |
| z_CERAD_IR | 0 | 0.0 |
| z_CERAD_DR | 0 | 0.0 |
| z_AFT | 0 | 0.0 |
| z_DSST | 0 | 0.0 |
| GlobalZ_raw | 0 | 0.0 |
| GlobalZ | 0 | 0.0 |
| FISI_z | 0 | 0.0 |
| DII_z | 0 | 0.0 |
| low_cognitive_performance | 0 | 0.0 |
| FISI_q | 0 | 0.0 |
| DII_q | 0 | 0.0 |

| Variable | Missing, n | Missing (%) |
|---|---|---|
| obese | 0 | 0.0 |

*Counts and percentages are unweighted and refer to the final analytic sample (n = 2455). Percentages are displayed to one decimal place; the underlying frozen counts are unchanged.*

**Figure S1. Survey-weighted receiver operating characteristic curves**

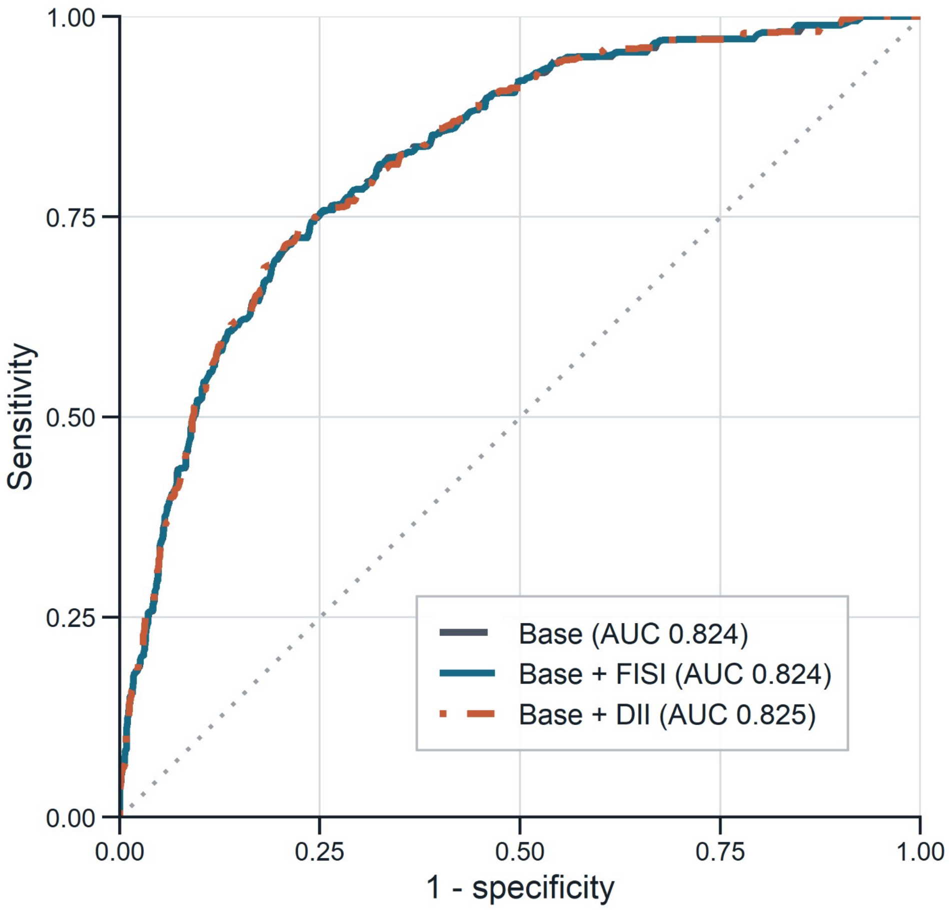


*Curves compare the fully adjusted base model with models adding FISI or DII. AUC, area under the curve; DII, Dietary Inflammatory Index; FISI, Food Inflammation Score for Individuals.*

**Figure S2. Sensitivity analyses for continuous global cognitive performance**

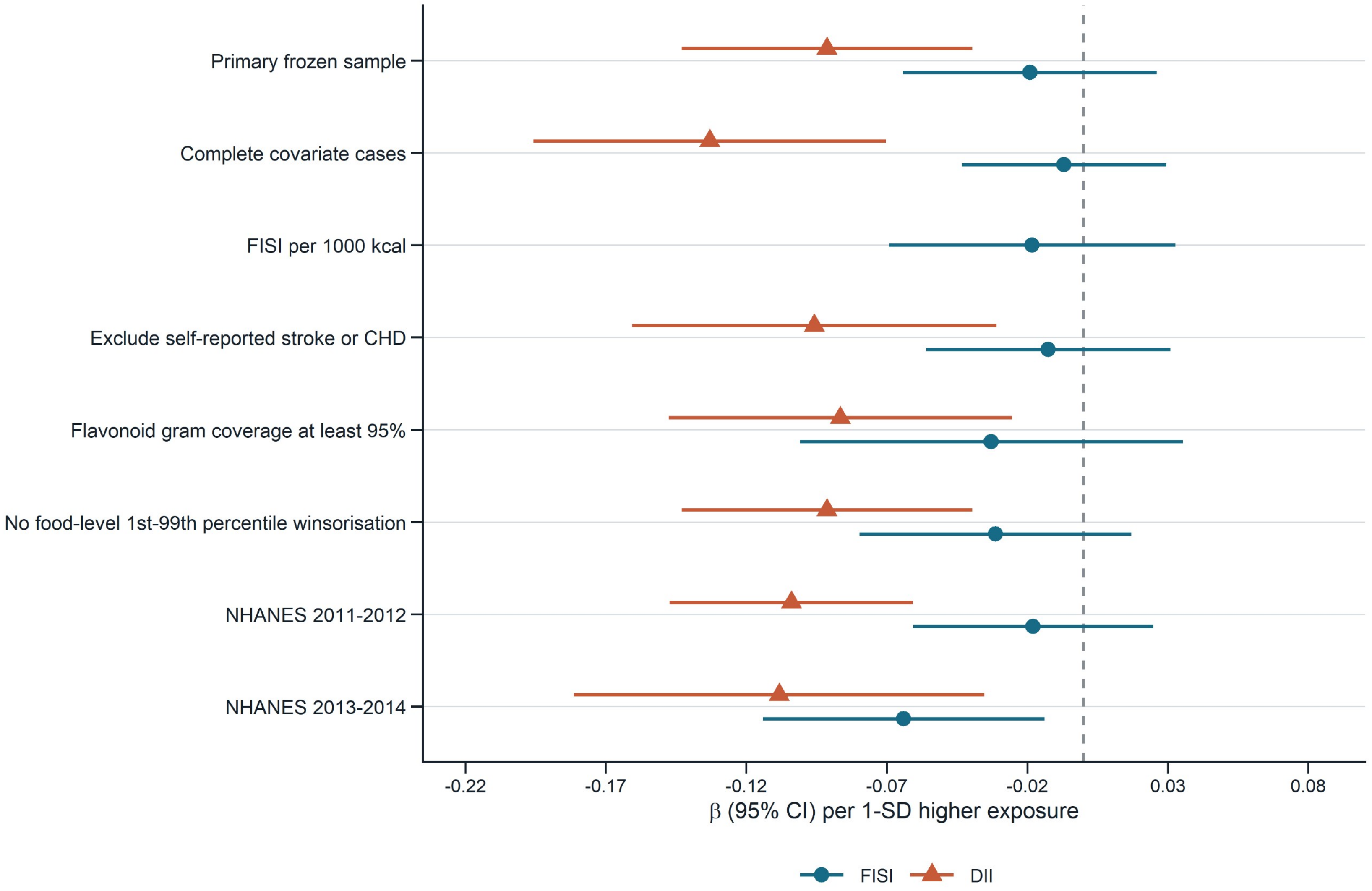


*Points are regression coefficients and horizontal bars are 95% confidence intervals per 1-SD higher exposure. Single-cycle estimates used normal-approximation Wald inference.*

#### Figure S3. Restricted cubic spline associations with global cognitive performance

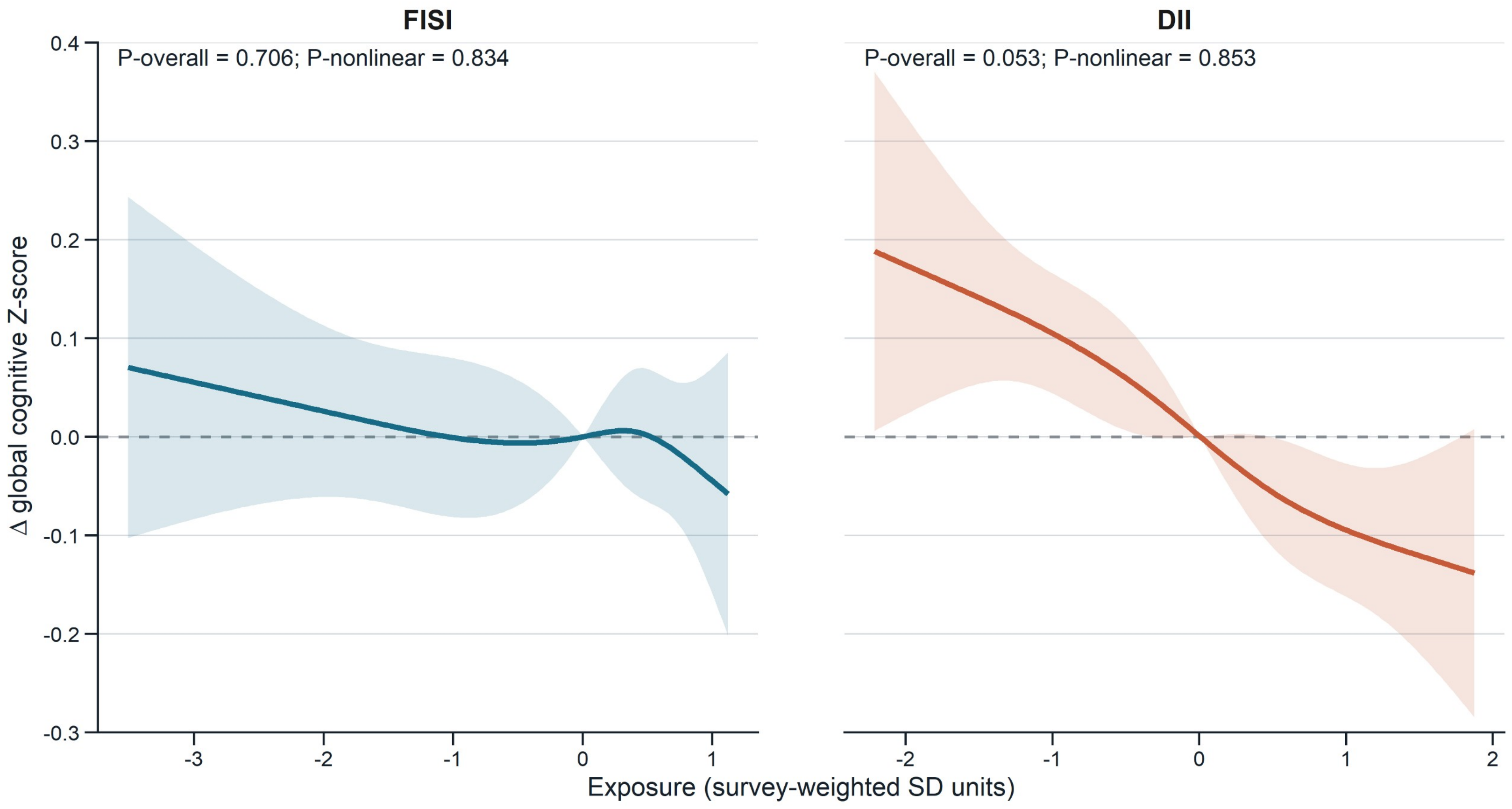


*Curves show fully adjusted differences in global cognitive Z-score; shaded bands are 95% confidence intervals. Exposures are expressed in survey-weighted SD units.*

### Figure S4. Survey-weighted calibration by decile of predicted probability

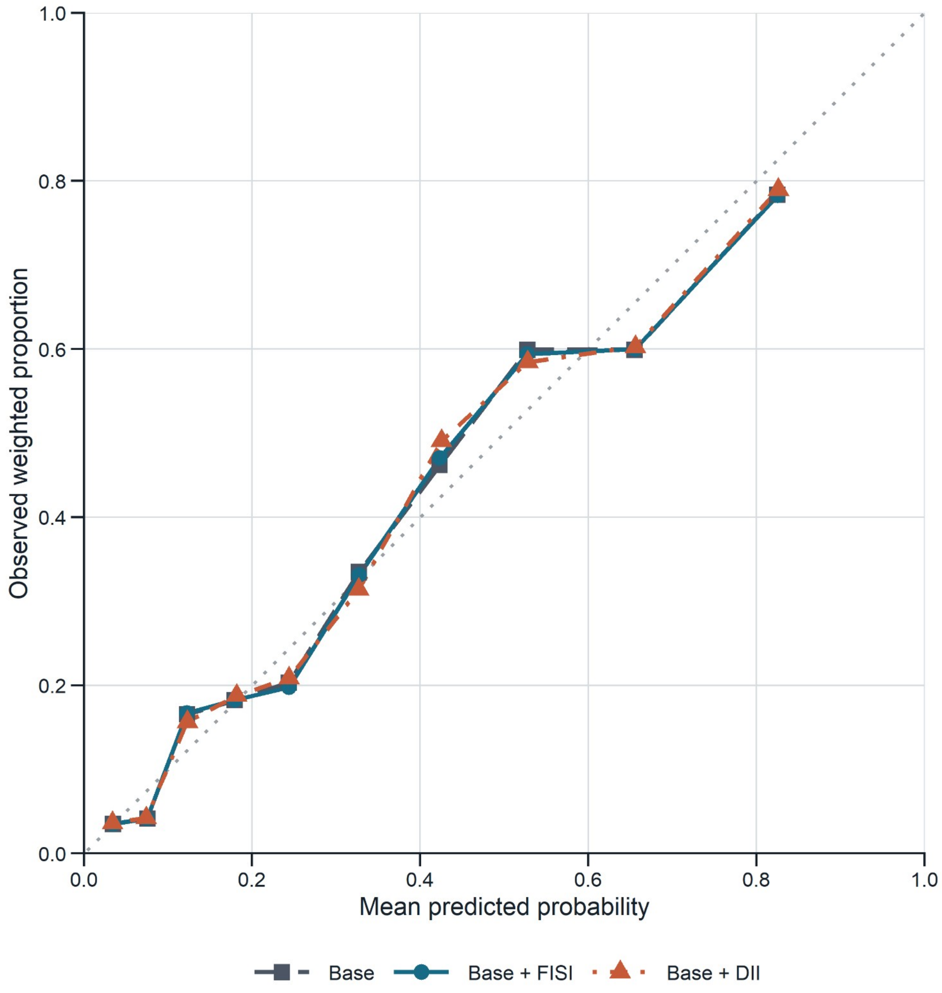


*Observed survey-weighted proportions are plotted against survey-weighted mean predicted probabilities for the base, base-plus-FISI, and base-plus-DII models. The diagonal is perfect agreement.*
